# The Impact of COVID-19 on Life Expectancy among Asian American Subgroups

**DOI:** 10.1101/2022.10.27.22281612

**Authors:** Sung S. Park, Noreen Goldman, Theresa Andrasfay

## Abstract

Despite recognition of the diversity of the Asian American population, the mortality impact of the COVID-19 pandemic has been limited to estimates for non-Hispanic Asian Americans in aggregate. This study provides estimates of life expectancy at birth before (2019) and during the pandemic, along with a broad set of demographic, health-related, and socioeconomic risk factors for COVID-19, for the six largest Asian American subgroups: Asian Indians, Chinese, Filipino, Japanese, Korean, and Vietnamese. Our study places these estimates in the context of the broader U.S. population by including the corresponding estimates for non-Hispanic Whites, non-Hispanic Blacks, Hispanics, and non-Hispanic Native Americans. We use data on age-specific all-cause mortality from CDC WONDER and population estimates from the 2015-2019 American Community Survey to construct life tables for each Asian subgroup by year. While losses in life expectancy during the second year of the pandemic diminished significantly among all racial/ethnic and Asian subgroups, these improvements do not compensate for the large increases in death rates in 2020. All major Asian subgroups except Japanese experienced greater losses of life in 2019-2020 as well as cumulatively (2019-2021) than Whites, with Vietnamese, Filipinos, and other South/Southeast Asians having suffered the largest declines in life expectancy among non-Hispanic Asians. Vietnamese and other Southeast Asians experienced the greatest cumulative losses across all racial/ethnic groups except Native Americans. Our findings underscore the heterogeneity of loss in life expectancy within the Asian American population while identifying some of the risk factors that likely underlie this large variation.

## Introduction

Over the past two years, the media and the research community have each written extensively on the COVID-19 pandemic, revealing the disparate rates of infection, hospitalization, and fatality across racial/ethnic groups in the U.S. Early reports promoted awareness of the disproportionate impacts of COVID-19 on the Black^*^ and Hispanic populations, both of whom suffered devastating declines in life expectancy that substantially exceeded those in the White population (1, 2). More recently, studies of Native Americans revealed their losses were even larger, surpassing those of all other major racial/ethnic groups (3, 4). However, as of October 2022, there remains a striking absence of research on COVID-related changes in life expectancy across the Asian American population, particularly by subgroup (5).

This shortcoming is consistent with a more general dearth of studies on the determinants and consequences of COVID-19 for Asian Americans (6), and a longer-term omission of Asian Americans in health-focused research (7, 8, 9). Two exceptions that explicitly examine Asian American life expectancy during the pandemic are a recent National Vital Statistics Systems (NVSS) report providing national estimates of life expectancy for 2021 (2), and a paper comparing life expectancy losses during the pandemic worldwide (10). Both studies calculated a substantial loss in life expectancy from 2019 through 2021 of about two years for Asian Americans (2, 10), and indicate a shrinking Asian American mortality advantage relative to Whites during the pandemic.

While these studies reflect progress in their inclusion of Asian Americans when examining pandemic-related health disparities, aggregated estimates of Asian Americans as a whole mask potential differences by subgroup. Scholars have established significant variability in longevity across the six largest Asian American subgroups: Asian Indians, Chinese, Filipinos, Japanese, Koreans, and Vietnamese. For example, during the period 2012-2016, life expectancy varied by as much as a decade across the six major Asian American subgroups from a low of 77.5 and 79.7 years among Vietnamese males and females, respectively, to a high of 86.8 and 91.3 among Chinese males and females, respectively, a range that greatly exceeds that between the White and Black populations (7). These disparities are the result of complex multifactorial etiology among subgroups in their demographic characteristics, migration histories, English fluency, socioeconomic integration (such as educational attainment and occupation), and experiences with discrimination (7, 11, 12).

Such heterogeneity makes it problematic for researchers to analyze Asian Americans as a single group. Regional studies have uncovered high rates of hospitalization and high case fatality rates (ratio of deaths to infections from COVID-19) among South/Southeast Asians and Chinese during the early part of the pandemic (8, 13). While population-level data are preferred, disaggregating the major subgroups at the national level requires that analysts deal with numerous data limitations: lack of information on specific subgroups in some sources, high rates of missing and misclassified ethnic identity in the data, and relatively small sample sizes (11). Although Xu (14) recently provided national mortality estimates for Asian American subgroups during the first year of the pandemic, his analysis relies on age-adjusted death rates, a metric that is less intuitive than life expectancy, particularly for comparisons across Asian American subgroups or between Asian Americans and other racial/ethnic groups. Life expectancy (typically calculated at birth) summarizes the mortality experience of a population at all ages during a specified year or period in a way that is independent of the population’s age distribution and indicates how long individuals could expect to live throughout their lifetimes under the mortality regime of the particular period.

In this paper, we fill an important gap in the pandemic literature by providing the first national estimates of life expectancy at birth (e_0_) prior to and during the pandemic and associated estimates of loss of years of life among the six largest, single-race Asian American groups in the U.S.: Asian Indians, Chinese, Filipinos, Japanese, Koreans, and Vietnamese. Although we focus our discussion on these six groups, we also include estimates for a heterogeneous residual category often referred to as “Other Asians.” Because almost 88% of this group identified with one of 13 South or Southeast Asian national origin groups, we refer to this category as “Other South/Southeast Asians”.^†^ We use data from CDC Wonder (2019, 2020 and 2021) and a five-year American Community Survey (ACS) sample (2015-2019) for our calculations, whilst providing survey-based national estimates of demographic, migration-related, health-related, and socioeconomic risk factors that likely contribute to variability in exposure to viral transmission and fatality.

## Results

### Profiles of Asian American Subgroups

Previous research has identified significant heterogeneity within the Asian American population on a broad range of demographic, migration-related, health, and socioeconomic characteristics (11, 16). Table 1 presents estimates of many of these measures for Asian American subgroups and, for comparative purposes, other major racial/ethnic groups. All groups, except the Hispanic population, include only non-Hispanics. Separate estimates by sex, where available, are shown in Appendix Table S1.

**Table 1.** Summary of Characteristics by Race/Ethnicity/Asian American Subgroup.

|  | NH<br>Asian [1] | Asian<br>Indian | Chinese | Filipino | Japanese | Korean | Vietnamese | Other<br>S/SE Asian | NH White | NH Black | Hispanic | NH AI/AN | Source(s) |
| --- | --- | --- | --- | --- | --- | --- | --- | --- | --- | --- | --- | --- | --- |
| **Demographic** |  |  |  |  |  |  |  |  |  |  |  |  |  |
| Mean Age | 38.0 | 34.0 | 39.2 | 41.5 | 49.3 | 40.3 | 38.3 | 33.0 | 42.2 | 35.9 | 31.5 | 35.7 | [6] |
| % 65+ | 12.5 | 8.3 | 13.9 | 15.9 | 27.6 | 14.0 | 12.0 | 7.5 | 19.8 | 11.4 | 7.3 | 11.3 | [6] |
| Mean HH Size | 3.0 | 3.0 | 2.8 | 3.4 | 2.3 | 2.7 | 3.4 | 3.7 | 2.4 | 2.6 | 3.6 | 2.9 | [6] |
| % in Multigenerational HH [2] | 38.5 | 30.6 | 35.3 | 49.8 | 26.6 | 30.6 | 48.6 | 45.0 | 25.2 | 38.8 | 42.4 | 40.1 | [6] |
| **Migration-Related** |  |  |  |  |  |  |  |  |  |  |  |  |  |
| % Foreign-Born | 66.8 | 71.4 | 68.6 | 65.7 | 41.0 | 70.5 | 66.6 | 63.3 | 4.0 | 9.5 | 34.4 | 1.2 | [6] |
| % Arrived U.S. < Five Years Ago (Foreign-Born) | 18.2 | 25.5 | 19.7 | 11.5 | 26.3 | 10.8 | 10.8 | 18.9 | 14.9 | 16.8 | 10.6 | 14.7 | [6] |
| % Limited Ability to Speak English (5+ and Foreign-Born) | 19.9 | 7.6 | 30.2 | 6.6 | 17.5 | 24.5 | 36.7 | 22.7 | 11.0 | 8.4 | 41.6 | 7.2 | [6] |
| **Health Conditions and Behaviors** |  |  |  |  |  |  |  |  |  |  |  |  |  |
| Self-Reported Obesity (Adjusted by Age and Sex) (18+) | 11.7 | 11.2 | 6.5 | 16.8 | 15.3 | 8.5 | 6.3 | N/A | 29.4 | 39.7 | 33.2 | 37.1 | [7], [8] |
| Self-Reported Diabetes (18+) | 8.4 | 10.3 | 4.7 | 14.0 | 12.8 | 4.0 | 3.2 | N/A | 10.1 | 14.7 | 11.8 | 16.1 | [7], [8] |
| Current Smoker (18+) | 7.6 | 5.5 | 6.0 | 8.9 | 9.4 | 13.4 | 8.4 | N/A | 17.0 | 18.3 | 13.2 | 29.9 | [7], [8] |
| % Without Health Insurance (25-64) | 8.2 | 5.6 | 7.8 | 6.7 | 4.5 | 11.7 | 10.0 | 12.5 | 8.8 | 15.1 | 26.6 | 25.4 | [6] |
| % No COVID-19 Vaccination By April 2021 (18+) | 30.4 | N/A | N/A | N/A | N/A | N/A | N/A | N/A | 41.0 | 53.7 | 52.7 | 61.3 | [9] |
| % No COVID-19 Vaccination By Nov. 2021 (18+) | 4.8 | 2.2 | 4.8 | 7.6 | 7.1 | 5.8 | 10.0 | 13.5 | 21.3 | 21.8 | 18.7 | 38.2 | [9] |
| **Socioeconomic** |  |  |  |  |  |  |  |  |  |  |  |  |  |
| % Completed College (25+) | 54.4 | 75.3 | 56.4 | 49.5 | 52.3 | 56.9 | 30.7 | 39.2 | 35.7 | 21.6 | 16.1 | 15.4 | [6] |
| Median HH Income (2019 \$) | 87,355 | 119,120 | 80,965 | 94,961 | 81,820 | 71,221 | 68,739 | 65,000 | 68,000 | 41,336 | 51,000 | 42,000 | [6] |
| % Self-Employed (25+ and Employed) | 10.1 | 8.8 | 10.5 | 5.4 | 11.3 | 17.7 | 14.0 | 9.9 | 11.7 | 5.6 | 9.9 | 7.7 | [6] |
| % Frontline Workers (25+ and Employed) [4] | 47.1 | 32.4 | 40.5 | 60.5 | 37.4 | 44.7 | 64.8 | 59.1 | 49.2 | 60.5 | 69.2 | 60.5 | [6] |
| % Commuting By Taking Public Transportation (25+, Employed, and Worked Last Week) [5] | 10.4 | 10.9 | 16.4 | 8.1 | 8.2 | 8.8 | 3.9 | 8.3 | 3.1 | 9.4 | 6.4 | 3.0 | [6] |
| N (Weighted) [6] | 17,653,671 | 3,967,780 | 4,347,702 | 2,818,434 | 780,787 | 1,466,175 | 1,844,814 | 2,427,979 | 197,097,542 | 39,973,076 | 55,689,921 | 2,151,718 |  |

Notes: Weighted using individual weights. Household income weighted using household weights. All Asian American subgroups consist of single-race, non-Hispanic individuals. NH: Non-Hispanic. S/SE: South or Southeast.
AI/AN: American Indian and Alaskan Native. N/A: Not available.
[1] Unless otherwise noted, NH Asian is comprised of: NH Asian Indian, NH Chinese, NH Filipino, NH Japanese, NH Korean, NH Vietnamese, and NH Other S/SE Asian individuals.
[2] Households with at least two adult generations or a skipped generation (grandparent-grandchild) are considered multigenerational.
[3] Does not speak English or speaks English but not well.
[4] Modes of public transportation are: bus, trolley bus, streetcar, light rail, subway, train, and ferryboat.
[5] Frontline occupations are defined using classification rules in Goldman et al. (2021).
[6] American Community Survey (2015-2019).
[7] Behavioral Risk Factor Surveillance System (BRFSS) 2013-2020 from Shah et al. (2022). NH Asian includes individuals in residual NH Asian categories.
[8] Hispanic and NH AI/AN estimates based on authors' calculations using 2013-2020 BRFSS.

The variables in Table 1 are potential risk factors for COVID-19 infection or severity that have previously been used to examine racial/ethnic disparities in COVID-19 outcomes (17, 18, 19). Viral transmission of the coronavirus occurs most frequently among older individuals (20), those living in large households (often multigenerational)^‡^, persons with limited English proficiency (who are most likely to be foreign-born), individuals with certain health conditions and co-morbidities (most notably obesity and diabetes), smokers, persons with poor access to health care (often due to lack of health insurance), individuals with low educational attainment and low income, frontline workers (who are unable to work remotely), those relying on public transport for work, and persons who did not receive any COVID-19 vaccination. Some of these factors – such as age, presence of other chronic conditions, access to health care, and receipt of vaccines – are also related to disease severity and fatality.

A comparison of the aggregated Asian American population (“NH Asian”) with the other major racial/ethnic groups indicates several ways in which Asian Americans are advantaged with respect to the risks of infection. They are less likely to smoke, be obese, or to hold frontline jobs, and more likely to have health insurance, be vaccinated against COVID-19, and have higher levels of educational attainment and income (21, 22). On the other hand, Asian Americans have several notable disadvantages with regard to risk of (severe) infection: they are more likely than other groups to live in large households (except for Hispanics), to have more limited English speaking abilities (except for Hispanics), and to use public transport for commuting to work (likely because they tend to live in cities) (23). However, these simple comparisons mask large differences within the Asian American population.

As shown in the first panel of Table 1, the Japanese population is considerably older than the other South/Southeast American subgroups – 15 years older, on average, than Asian Indians. This differential is related to Japanese having a higher proportion of individuals who are native born rather than (recent, younger) immigrants relative to other groups (see “% Foreign-born” in Table 1). Japanese also have the lowest percentage living in multigenerational households among Asian American subgroups. In contrast, Filipinos, Vietnamese, and Other South/Southeast Asians are the most likely to live in a multigenerational household across all racial/ethnic groups.

Limited proficiency in English hampers access to health care, government services, and rapidly changing information about the transmission and mitigation strategies for newly identified infectious diseases like COVID-19 (24). The prevalence of limited English-speaking ability varies widely across foreign-born Asian American subgroups from under 7% for Filipinos to 37% among Vietnamese and only slightly lower for Chinese. However, we note that English proficiency is not necessarily highly correlated with the percent foreign-born or the recency of immigration across subgroups, as English is one of the official languages in India and the Philippines.

Few researchers have provided recent national estimates of health conditions for the largest six Asian American subgroups. Health information based on national samples generally comes from modest-sized sample surveys, often requiring access to restricted data on specific Asian ethnicity as well as pooling of data across survey years to achieve stable estimates for small subgroups. The third panel of Table 1 presents the prevalence of obesity and diabetes, two important health-related risk factors for COVID-19 infection and severity (25), along with data on current smoking, based on estimates from the Behavioral Risk Factor Surveillance Survey (BRFSS) for 2013-2020. These values are conservative because they are based on self-reports (of height, weight, and diabetes) and low response rates (45% to 49% across years) that likely reflect selective inclusion of respondents of high socioeconomic status. We do not present estimates for cardiovascular conditions because researchers have shown these self-reports to be considerably more biased than those for diabetes (26).

The results for obesity and diabetes in Table 1 are consistent with earlier studies that highlight immense variation in cardiometabolic risk factors across Asian American subgroups (27). The estimated prevalence of diabetes varies almost fivefold, from about 3% among Vietnamese to about 14% among Filipinos, with estimates for Filipinos and Japanese exceeding those for the White and Hispanic populations. The estimates of obesity (BMI cutoff of 30) also vary considerably among Asian American subgroups, but all are far below those for the other major racial/ethnic groups. However, studies have indicated that Asian Americans are at risk for deleterious cardiometabolic outcomes (including diabetes) at lower BMI cutoffs than some other populations. The WHO has suggested a BMI cutoff of 27.5 for obesity among Asian Americans (27, 28), a value that would roughly double the prevalence in Table 1. Although generally lower among Asian Americans than other groups, the prevalence of smoking among Koreans (13%) is similar to that of Hispanics but still below frequencies for the White, Black, and Native American populations.

An important component of access to health care is possession of health insurance, which not only affects the likelihood of healthcare and treatment during the pandemic but also receipt of vaccination. The percentage of Asian Americans (ages 25-64) without health insurance (8%) is lower than corresponding values for other groups, but again varies substantially from a low of about 5% among Japanese to at least 10% among Vietnamese and Koreans. Since insurance is often provided by employers, some of this variation results from differential rates of self-employment (29): among workers, Vietnamese and Koreans have the highest rate of self-employment, as shown in the fourth panel of the table. Furthermore, an effective health behavior among the Asian American population during the pandemic has been earlier and more extensive vaccination: according to self-reported information from the National Immunization Survey Adult COVID Module (30), by April 2021, only 30% of Asian Americans did not receive any vaccine, compared with 41% to 61% among other major racial/ethnic groups. By November 2021, fewer than 5% of Asian Americans had received no vaccination, a value that varies between 2% among Asian Indians and 10% among Vietnamese, values that continue to be far below those for the other racial/ethnic groups.

Socioeconomic status varies substantially across Asian American subgroups, whether measured by educational attainment, income, or type of employment. The most dissimilar groups are Vietnamese and Asian Indians. Vietnamese adults (25+) are less than half as likely to have completed a college education as Asian Indians (31% versus 75%), about twice as likely to have frontline occupations among the working population (65% versus 32%), and have median household incomes that are $50k less ($69K versus $119K). These differences reflect the occupational segregation of these subgroups, with Vietnamese workers frequently employed in personal care and service occupations (e.g., the nail salon industry), in contrast to Asian Indian workers who often work in computing and consulting industries (31). Potential exposure to infection via public transport also varies considerably across Asian American subgroups, from only about 4% among Vietnamese who worked in the past week to about 16% among Chinese, the latter of whom have the lowest rates of suburbanization of all Asian American subgroups (23).

### Loss of Life from 2019 to 2021

Table 2 provides estimates of life expectancy at birth for 2019, 2020, and 2021 with 95% confidence intervals, annual losses (2019-2020 and 2020-2021), and the two-year decline (2019-2021), by sex and race/ethnicity/Asian American subgroup. Both before and during the pandemic, Asian Americans as a whole, as well as each of the six major subgroups, had higher life expectancy than each of the other racial/ethnic groups. Pre-pandemic life expectancy values among the six Asian American subgroups ranged from 85.5 among Asian Indians to 89.2 among Chinese; Filipinos had the lowest life expectancy among males (82.4) and Asian Indians had the lowest value among females (87.4), whereas Chinese had the highest estimates for males (87.0) and females (91.2). ^§^

**Table 2.**
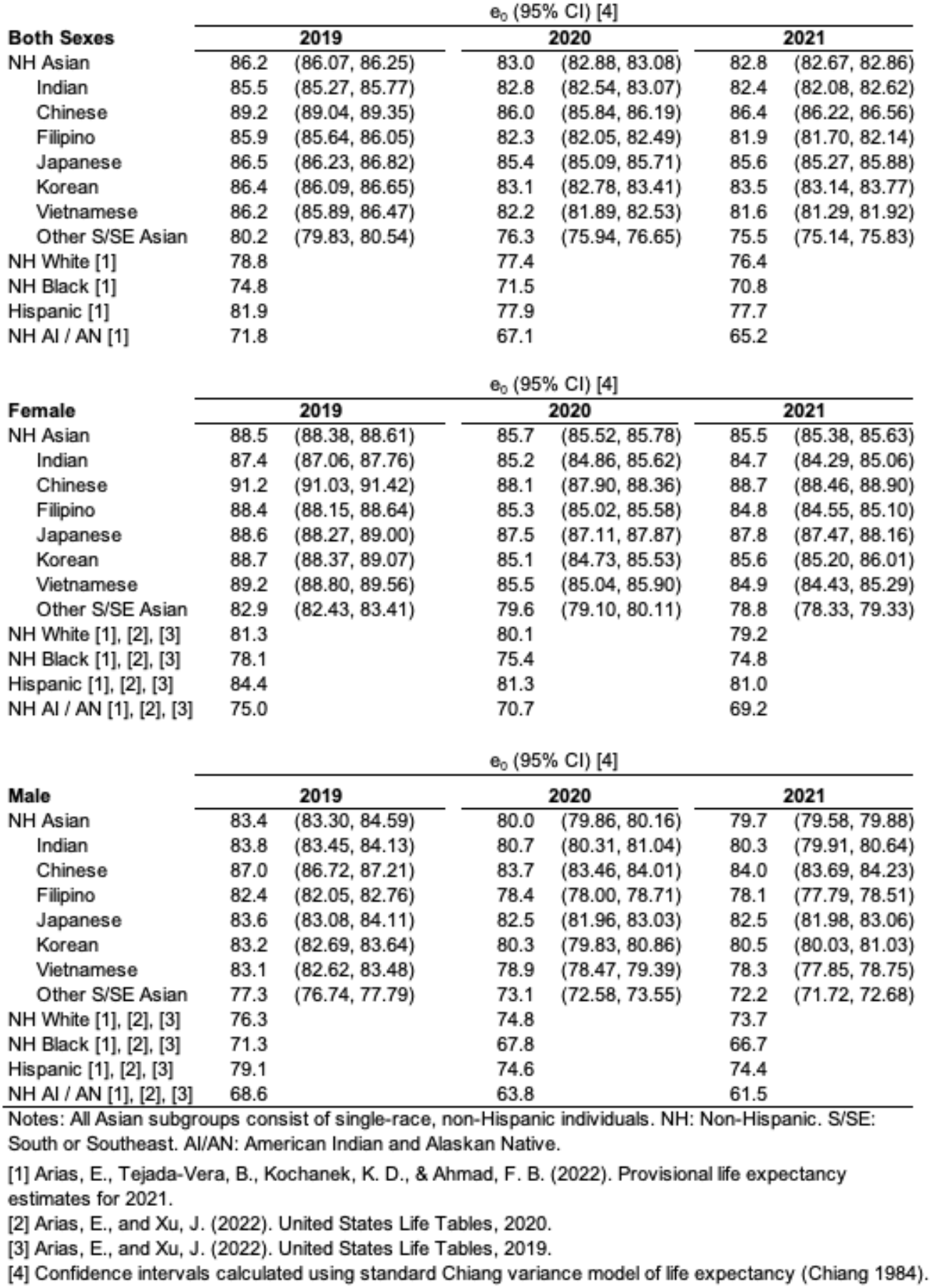
Life Expectancy at Birth by Race/Ethnicity/Asian American Subgroup, Year, and Sex.

In Figure 1, we depict these estimates for Asian American subgroups for each year, along with changes in life expectancy (Δ) year over year (green numbers reflecting 2019-2020 change, blue numbers reflecting 2020-2021 change, and black numbers reflecting 2019-2021 change). As shown in the top panel of the figure (“Both Sexes”), losses during the first year of the pandemic for the Asian American population were large, 3.2 years on average – a decline almost one year larger than for Whites – with the loss exceeding three years for four of the six major Asian American subgroups. Filipinos, Other South/Southeast Asians and Vietnamese, suffered the greatest declines in 2020 (3.6, 3.9, and 4.0 years, respectively), on par with the large loss experienced by the Hispanic population; in contrast, Japanese experienced a loss of just over one year.

**Figure 1.**
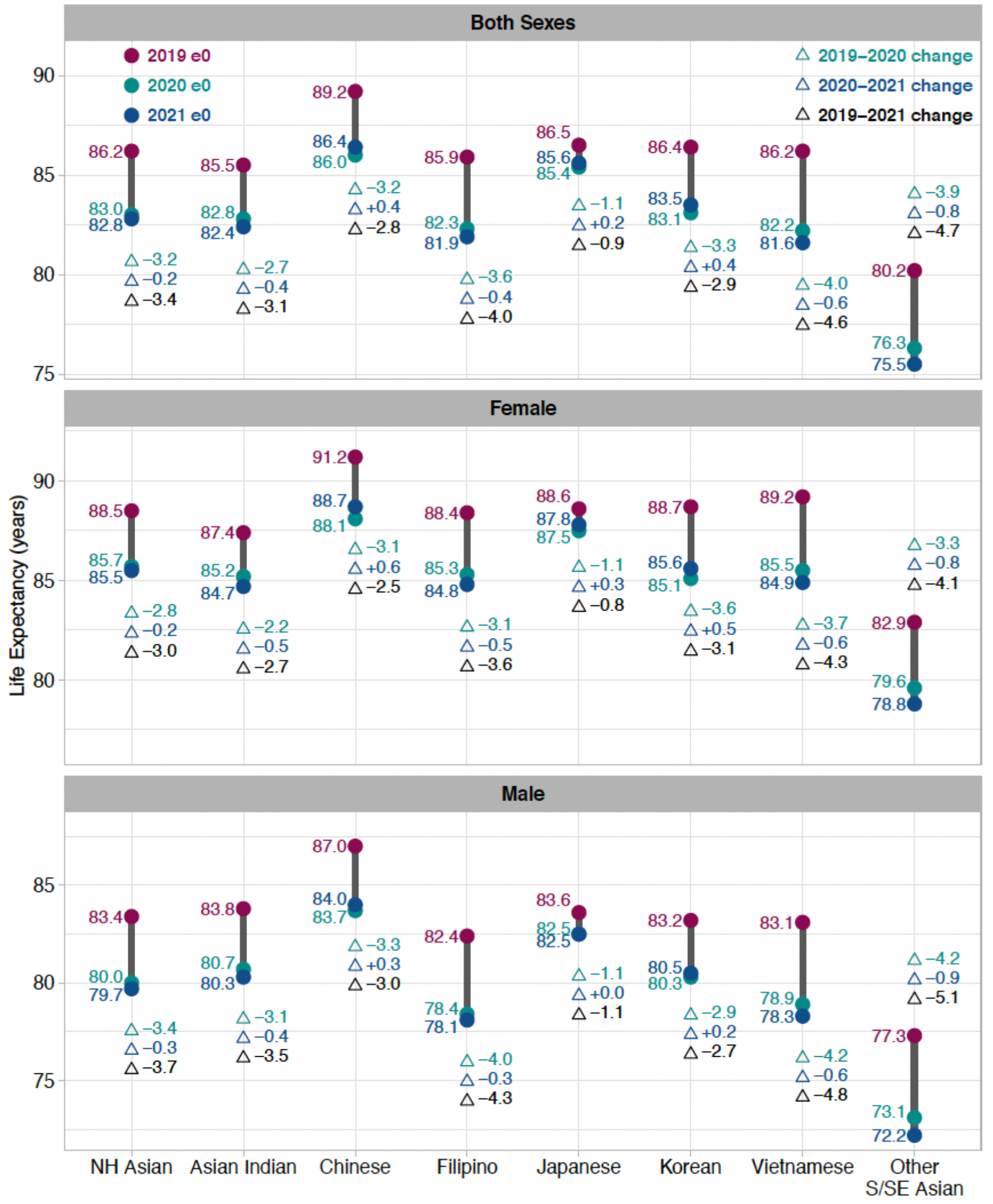
Life Expectancy at Birth and Loss of Life from 2019-2021 by Race/Ethnicity/Asian Subgroup and Sex.

The average loss of life in 2021 relative to 2020 was modest among Asian Americans (0.2 years as shown in the top panel of Figure 1), reflecting both losses and gains across subgroups with an overall decline considerably below those of the White and Native American populations. For Chinese, Japanese, and Koreans, life expectancy recovered modestly in 2021. Nevertheless, because of the large drops in 2020, life expectancy in 2021 remains considerably below values prior to the pandemic, especially among Filipinos and Vietnamese. Filipinos’ cumulative loss (4.0 years) is similar to those of the Black and Hispanic populations, while the cumulative loss (4.6 years) for Vietnamese exceeds those for each group presented in Table 2 except Native Americans (6.6 years). In all Asian American and non-Asian American groups except Koreans, male losses over the period 2019-2021 exceeded those of females.

## Discussion

Despite intensified calls to improve data collection systems and reporting of health measures across Asian American subgroups (35), studies of the impact of the COVID-19 pandemic on life expectancy by subgroup have been noticeably absent. Yet, there is substantial variation across the Asian American population because of differences in the same types of risk factors for exposure to and severity of infection that have previously been identified in the other major racial/ethnic groups in the U.S. (17, 36). Occupational exposure, pre-existing health conditions, living arrangements, and vaccination uptake have contributed to greater losses among Blacks, Hispanics, and Native Americans compared to their White counterparts (36, 37, 38). Our study takes advantage of recently released mortality data for the six largest Asian American subgroups (Asian Indians, Chinese, Filipinos, Japanese, Koreans, and Vietnamese) by the NVSS to examine losses in life expectancy at birth before and during the COVID-19 pandemic for each subgroup, and contextualizes the differences compared to other racial/ethnic groups that have been analyzed to date.

In 2020, compared to the pre-pandemic period 2019, Asian Americans as a whole suffered greater losses than Whites but lower losses than Black, Hispanic, and Native American populations. Yet, the aggregate Asian American estimate of the loss of life expectancy masks significant within-group variation, particularly between East Asian and non-East Asian subgroups. Japanese losses were smallest at just over one year, whereas Chinese and Koreans experienced declines comparable to the losses among Blacks (3.3 years). Filipino, Vietnamese, and Other South/Southeast Asian losses of life were even greater (3.6 to 4.0 years), similar to the losses among Hispanics during the same time period (4.0 years). Despite higher pre-pandemic life expectancies among Asian Americans than other racial/ethnic groups, this advantage is not strongly associated with the magnitude of loss experienced during the first year of the pandemic. This finding contradicts our expectation that a relatively high socioeconomic status and good health would have translated to greater protective effects against disease transmission and severity.

Although the risk factors summarized in Table 1 likely account for a substantial part of the observed losses of life expectancy across groups, a statistical decomposition of the relative importance of these factors is not feasible given six groups for comparison. Nevertheless, we note both shared and contrasting characteristics for Vietnamese and Filipinos, the two groups with the largest losses in 2019-2020 among Asian American subgroups. These two groups have the largest mean household sizes and are the most likely to be working frontline jobs. However, there are two other distinct risk factors that likely explain the large loss of life among Vietnamese. Their lower levels of English proficiency likely limited information about COVID-19 transmission and reduced utilization of medical treatment during the pandemic. Furthermore, high levels of self-employment among Vietnamese may have required some to continue working in high viral exposure jobs such as positions in personal service industries (31). Filipinos, on the other hand, have high rates of two important risk factors for COVID-19 infection and severity: obesity and diabetes. Although many of their frontline jobs are relatively higher-status positions in the medical field (39) in contrast to Vietnamese occupations, there were numerous reports of limited access to mitigation strategies such as personal protective equipment for Filipino healthcare workers during the early stages of the pandemic (40).

Loss of life during the second year of the pandemic decreased substantially. In 2020-2021, all Asian American subgroups suffered a smaller loss than in 2019-2020, smaller than all other racial/ethnic groups except Hispanics. Furthermore, several groups (Chinese, Japanese, and Koreans) made modest recoveries, which may be partially attributable to higher vaccination and booster rates (41). Although data on mitigation behaviors by subgroup are limited, Asian Americans have been noted elsewhere as having higher rates of handwashing, isolation, and masking than other groups (6).

The relative differences in the cumulative (2019-2021) losses from COVID-19 among Asian American subgroups principally reflect the variation in increased mortality during the first year of the pandemic. Based on the provisional death counts for 2021, it appears that all subgroups, with the exception of Japanese, experienced greater overall losses of life compared to Whites. Moreover, Vietnamese and Other South/Southeast Asians experienced greater losses of life than Blacks and Hispanics but not Native Americans, who suffered the largest mortality impact among all groups. Only through disaggregation are we able to identify these stark disparities among South Asians and Southeast Asians compared to East Asian subgroups.

Our findings underscore the heterogeneity of loss in life expectancy among the Asian American population and the importance of further efforts to examine subgroup differences, particularly among Filipinos, Vietnamese, and Other South/Southeast Asian subgroups that have suffered the most severe loss of life during the pandemic. We recognize, however, that researchers – including ourselves – are limited by the data that are currently available. For example, we relied on five-year data from the ACS for population estimates by subgroup because the Census Bureau’s postcensal population estimates are unavailable at the Asian American subgroup level. Furthermore, despite varying immigration histories and processes of integration among South Asians and Southeast Asians, we were limited to grouping these individuals into a single “Other South/Southeast Asian” category due to small sample size constraints. Native Hawaiian and Pacific Islanders, distinct from Asian Americans, warrant their own analysis by subgroup (42). Finally, while the psychological impact of increased anti-Asian violence during the pandemic has been well-documented (11), its implications for COVID-19 transmission and infection through changes in healthcare access, employment, and social distancing are limited and not well-understood (43). We encourage scholars to continue efforts to include Asian American subgroups in studies of both the social determinants and magnitude of health disparities across the U.S. population.

## Materials and Methods

### Statistical Analysis

Descriptive statistics for select characteristics in Table 1 were computed using STATA 16.1. To obtain life expectancy at birth for each racial/ethnic group and Asian American subgroup, by sex, we created abridged life tables for 2019, 2020, and 2021 based on all-cause mortality data with 10-year age intervals except for the following three age groups: infants dying before age 1, 1-14 years, and the open-ended age interval for individuals 85 years and older. Death counts obtained through CDC WONDER (44) for 2019 and 2020 are considered final, while 2021 deaths are provisional and subject to change. Population counts were obtained from the 2015-2019 ACS (45). We adjusted death counts for 2019, 2020, and 2021 for racial misclassification using the sex-and age-specific misclassification ratios published by Arias et al. (2). To compute 1a0 (average person years in the first year of life) for each Asian American subgroup by sex, we used Preston et al.’s (46) formulae adapted from Coale and Demeny (47). Estimates of life expectancy at birth are based on standard life table approaches (46). 95% confidence intervals for life expectancy estimates were calculated using Chiang’s approach (48). All life expectancy and confidence interval calculations were computed using Microsoft Excel.

## Data Availability

All data produced in the present work are contained in the manuscript.

## Data Availability

Population and death counts for each Asian American subgroup by sex and year obtained from the ACS and CDC WONDER, respectively, are provided in Appendix Table S2. Characteristics for select measures in Table 1 may be obtained using publicly available data from IPUMS USA (usa.ipums.org) and the BRFSS website (https://www.cdc.gov/brfss/index.html) (49).

## Acknowledgments

The authors thank Keunbok Lee for providing code used to identify frontline workers in this study. S.S.P. acknowledges support from the National Institute on Aging (T32-AG033533). T.A. acknowledges support from the National Institute on Aging (Award Number T32AG000037). There was no specific funding source for this study.

## Appendix

**Table S1.** Summary of Characteristics by Race/Ethnicity/Asian American Subgroup and Sex.

|  | NH<br>Asian [1] | Asian<br>Indian | Chinese | Filipino | Japanese | Korean | Vietnamese | Other<br>S/SE Asian | NH White | NH Black | Hispanic | NH AI/AN |
| --- | --- | --- | --- | --- | --- | --- | --- | --- | --- | --- | --- | --- |
| **Demographic** |  |  |  |  |  |  |  |  |  |  |  |  |
| Mean Age |  |  |  |  |  |  |  |  |  |  |  |  |
| Female | 39.1 | 33.9 | 39.8 | 43.4 | 51.4 | 42.2 | 38.9 | 33.6 | 43.2 | 37.2 | 32.1 | 36.6 |
| Male | 36.8 | 34.1 | 38.5 | 39.1 | 46.6 | 37.8 | 37.6 | 32.3 | 41.1 | 34.4 | 30.8 | 34.8 |
| % 65+ |  |  |  |  |  |  |  |  |  |  |  |  |
| Female | 13.6 | 8.4 | 14.3 | 17.6 | 30.8 | 15.9 | 12.7 | 8.2 | 21.5 | 13.0 | 8.3 | 12.2 |
| Male | 11.3 | 8.2 | 13.6 | 13.7 | 23.4 | 11.7 | 11.2 | 6.7 | 18.1 | 9.7 | 6.2 | 10.3 |
| **Migration-Related** |  |  |  |  |  |  |  |  |  |  |  |  |
| % Foreign-Born |  |  |  |  |  |  |  |  |  |  |  |  |
| Female | 69.0 | 71.2 | 71.2 | 70.3 | 47.4 | 73.7 | 68.9 | 64.7 | 4.1 | 9.5 | 34.3 | 1.2 |
| Male | 64.3 | 71.6 | 65.7 | 59.8 | 32.2 | 66.5 | 64.1 | 61.8 | 3.9 | 9.5 | 34.5 | 1.1 |
| % Arrived U.S. < Five Years Ago (Foreign-Born) |  |  |  |  |  |  |  |  |  |  |  |  |
| Female | 17.6 | 25.8 | 19.0 | 11.7 | 20.3 | 10.2 | 11.7 | 19.0 | 14.0 | 16.6 | 10.4 | 14.1 |
| Male | 18.8 | 25.3 | 20.6 | 11.1 | 38.3 | 11.7 | 9.7 | 18.8 | 15.8 | 17.1 | 10.7 | 15.4 |
| % Limited Ability to Speak English (5+ and Foreign-Born) [3] |  |  |  |  |  |  |  |  |  |  |  |  |
| Female | 21.8 | 10.3 | 30.9 | 6.4 | 16.4 | 26.5 | 40.5 | 26.4 | 12.3 | 10.1 | 44.7 | 6.5 |
| Male | 17.7 | 5.1 | 29.2 | 6.8 | 19.7 | 21.8 | 32.2 | 18.8 | 9.5 | 6.6 | 38.6 | 7.9 |
| **Health Conditions and Behaviors** |  |  |  |  |  |  |  |  |  |  |  |  |
| % Without Health Insurance (25-64) |  |  |  |  |  |  |  |  |  |  |  |  |
| Female | 7.7 | 5.5 | 7.4 | 6.5 | 4.0 | 10.7 | 9.5 | 11.3 | 7.5 | 11.6 | 23.5 | 21.8 |
| Male | 8.6 | 5.7 | 8.3 | 7.1 | 5.1 | 12.9 | 10.5 | 13.7 | 10.1 | 19.0 | 29.6 | 29.2 |
| **Socioeconomic** |  |  |  |  |  |  |  |  |  |  |  |  |
| % Completed College (25+) |  |  |  |  |  |  |  |  |  |  |  |  |
| Female | 52.3 | 71.8 | 54.4 | 52.1 | 48.5 | 53.1 | 29.2 | 37.3 | 35.8 | 23.9 | 17.6 | 16.8 |
| Male | 56.9 | 78.5 | 58.8 | 45.6 | 57.9 | 62.3 | 32.4 | 41.2 | 35.6 | 18.8 | 14.6 | 14.0 |
| % Self-Employed (25+ and Employed) |  |  |  |  |  |  |  |  |  |  |  |  |
| Female | 8.9 | 7.4 | 9.4 | 4.9 | 10.8 | 14.9 | 13.8 | 7.8 | 9.1 | 4.1 | 8.6 | 5.8 |
| Male | 11.2 | 9.7 | 11.6 | 6.0 | 11.8 | 20.5 | 14.3 | 11.6 | 14.0 | 7.4 | 10.9 | 9.6 |
| % Frontline Workers (25+ and Employed) [4] |  |  |  |  |  |  |  |  |  |  |  |  |
| Female | 49.4 | 38.2 | 41.0 | 60.4 | 34.8 | 46.5 | 67.1 | 57.3 | 41.6 | 53.2 | 58.9 | 51.1 |
| Male | 45.0 | 28.8 | 40.0 | 60.5 | 40.0 | 42.8 | 62.3 | 60.5 | 55.8 | 68.9 | 76.9 | 70.1 |
| % Commuting By Taking Public Transportation<br>(25+, Employed, and Worked Last Week) [5] |  |  |  |  |  |  |  |  |  |  |  |  |
| Female | 11.1 | 10.8 | 18.4 | 8.8 | 8.7 | 9.6 | 4.2 | 8.1 | 3.0 | 10.0 | 7.5 | 2.8 |
| Male | 9.9 | 11.0 | 14.5 | 7.2 | 7.7 | 8.1 | 3.6 | 8.5 | 3.2 | 8.6 | 5.6 | 3.2 |
| **N (Weighted)** |  |  |  |  |  |  |  |  |  |  |  |  |
| Female | 9,281,312 | 1,902,150 | 2,325,149 | 1,590,831 | 448,964 | 812,402 | 960,940 | 1,240,876 | 99,910,932 | 20,877,675 | 27,563,319 | 1,097,586 |
| Male | 8,372,359 | 2,065,630 | 2,022,553 | 1,227,603 | 331,823 | 653,773 | 883,874 | 1,187,103 | 97,186,610 | 19,095,401 | 28,126,602 | 1,054,132 |

Notes: Data from American Community Survey (2015-2019). Weighted using individual weights. All Asian American subgroups consist of single-race, non-Hispanic individuals. NH: Non-Hispanic. S/SE: South or Southeast. AI/AN: American Indian and Alaskan Native.
[1] Unless otherwise noted, NH Asian is comprised of: NH Asian Indian, NH Chinese, NH Filipino, NH Japanese, NH Korean, NH Vietnamese, and NH Other S/SE Asian individuals.
[2] Households with at least two adult generations or a skipped generation household considered multigenerational.
[3] Does not speak English or speaks English but not well.
[4] Modes of public transportation are: bus, trolley bus, streetcar, light rail, subway, train, and ferryboat.
[5] Frontline occupations are defined using classification rules in Goldman et al. (2021).

**Table S2.** Population and Death Counts by Race/Ethnicity/Asian American Subgroup, Year, and Sex.

| **Both Sexes** |  |  |  |  |
| --- | --- | --- | --- | --- |
|  | **Population** | **Deaths** |  |  |
|  | 2019-2021 | 2019 | 2020 | 2021 |
| NH Asian [1] | 17,653,671 | 70,530 | 91,174 | 92,430 |
| Asian Indian | 3,967,780 | 9,911 | 12,490 | 12,967 |
| Chinese | 4,347,702 | 15,908 | 20,591 | 19,898 |
| Filipino | 2,818,434 | 13,521 | 17,983 | 18,579 |
| Japanese | 780,787 | 8,619 | 9,507 | 9,358 |
| Korean | 1,466,175 | 6,287 | 8,339 | 8,023 |
| Vietnamese | 1,844,814 | 6,958 | 9,349 | 9,913 |
| Other S/SE Asian | 2,427,979 | 9,326 | 12,915 | 13,692 |

| **Female** |  |  |  |  |
| --- | --- | --- | --- | --- |
|  | **Population** | **Deaths** |  |  |
|  | 2019-2021 | 2019 | 2020 | 2021 |
| NH Asian [1] | 9,281,312 | 34,616 | 43,476 | 44,046 |
| Asian Indian | 1,902,150 | 4,039 | 4,904 | 5,182 |
| Chinese | 2,325,149 | 7,757 | 9,881 | 9,468 |
| Filipino | 1,590,831 | 7,086 | 8,955 | 9,373 |
| Japanese | 448,964 | 5,309 | 5,850 | 5,723 |
| Korean | 812,402 | 3,357 | 4,491 | 4,307 |
| Vietnamese | 960,940 | 2,891 | 3,861 | 4,124 |
| Other S/SE Asian | 1,240,876 | 4,177 | 5,534 | 5,869 |

| **Male** |  |  |  |  |
| --- | --- | --- | --- | --- |
|  | **Population** | **Deaths** |  |  |
|  | 2019-2021 | 2019 | 2020 | 2021 |
| NH Asian [1] | 8,372,359 | 35,914 | 47,698 | 48,384 |
| Asian Indian | 2,065,630 | 5,872 | 7,586 | 7,785 |
| Chinese | 2,022,553 | 8,151 | 10,710 | 10,430 |
| Filipino | 1,227,603 | 6,435 | 9,028 | 9,206 |
| Japanese | 331,823 | 3,310 | 3,657 | 3,635 |
| Korean | 653,773 | 2,930 | 3,848 | 3,716 |
| Vietnamese | 883,874 | 4,067 | 5,488 | 5,789 |
| Other S/SE Asian | 1,187,103 | 5,149 | 7,381 | 7,823 |

Notes: Population counts are weighted counts from American Community Survey 2015-2019. Death counts are from CDC WONDER. NH: Non-Hispanic. S/SE: South or Southeast. AI/AN: American Indian and Alaskan Native. NH Asian is comprised of: NH Asian Indian, NH Chinese, NH Filipino, NH Japanese, NH Korean, NH Vietnamese, and NH Other S/SE Asian individuals.
[1] All Asian subgroups consist of single-race, non-Hispanic individuals.

## Footnotes

* For brevity, we refer to non-Hispanic White, non-Hispanic Black, non-Hispanic Native American, and non-Hispanic Asian American groups as White, Black, Native American, and Asian American, respectively. All Asian subgroups are non-Hispanic.

† In descending order, these 13 national origin groups are: Pakistani (20.0%), Hmong (11.9%), Cambodian (10.4%), Thai (8.5%), Laotian (7.9%), Bangladeshi (7.6%), Burmese (7.0%), Nepalese (6.8%), Indonesian (3.1%), Sri Lankan (2.0%), Bhutanese (1.1%), Mongolian (0.8%), and Malaysian (0.8%). Asian Indians, Bangladeshis, Bhutanese, Malaysians, Nepalese, Pakistanis, and Sri Lankans are classified by the Census as South Asian, while Burmese, Cambodians, Hmong, Indonesians, Filipinos, Laotians, Malaysians, Thais, and Vietnamese are classified as Southeast Asians (15). The remaining 12% of the group identified with at least one Asian national origin group that is non-specified in the public use data.

† A household with more than two adult generations or a skipped generation (grandparent-grandchild) is treated as multigenerational.

§; While population counts for non-Hispanic Asian Americans as whole are available in CDC WONDER, population counts for each Asian American subgroup are unavailable (32). We addressed this data constraint by using the ACS data for population estimates for each Asian American subgroup and the aggregated non-Hispanic Asian American group. In comparing our life expectancy estimates for the non-Hispanic Asian American group to those produced in the NVSS Reports (2, 33, 34), we find differences ranging from −0.7 to 0.6 years, across the three years. When we use comparable population counts for the non-Hispanic Asian American group as the CDC, these differences range from −0.1 to 0.2 years.

